# Real-world experience with tofacitinib in ulcerative colitis - a systematic review and meta-analysis

**DOI:** 10.1101/2021.04.27.21256170

**Authors:** L Lucaciu, N Constantine-Cooke, N Plevris, S Siakavellas, LAAP Derikx, GR Jones, CW Lees

**Affiliations:** Edinburgh IBD Unit, Western General Hospital, Edinburgh, Scotland, UK; Iuliu Hatieganu University of Medicine and Pharmacy, Cluj-Napoca, Romania; MRC Human Genetics Unit, Institute of Genetics and Molecular Medicine, University of Edinburgh, Western General Hospital, Edinburgh, Scotland, UK; Inflammatory Bowel Disease Centre, Department of Gastroenterology and Hepatology, Radboud University Medical Centre, Nijmegen, the Netherlands; Centre for Inflammation Research, The Queen’s Medical Research Institute, University of Edinburgh, Scotland, UK; Centre for Genomics and Experimental Medicine, Institute of Genetics and Molecular Medicine, University of Edinburgh, Western General Hospital, Edinburgh, Scotland, UK

**Keywords:** JAKi, IBD, clinical trials, new therapie s

## Abstract

**Background and aims:** Evidence on the outcomes of tofacitinib therapy in real world ulcerative colitis (UC) patients is needed, as a number of these patients would not match the inclusion criteria for clinical trials. We have therefore summarised data derived from observational, real-world evidence (RWE) studies on the effectiveness and safety of tofacitinib in moderate to severe ulcerative colitis (UC) patients.

**Methods:** We searched the PubMed, EMBASE, Scopus, Web of Science and Cochrane databases for observational studies on the use of tofacitinib in UC patients, published between 30/05/2018 and 24/01/2021. Pooled induction (8-14 weeks) and maintenance (16-26 weeks) clinical response and remission rates were calculated, as well as the proportion of reported adverse events using random effects models.

**Results:** Nine studies were included, comprising 830 patients, of which 81% were previously treated with anti-TNF and 57% with vedolizumab. Induction of clinical response and remission were achieved in 51% (95% CI 41-60%) and 37% (26-45%) of patients, after a median follow-up of 8 weeks. At the end of a median follow-up of 24 weeks, maintenance of clinical response and remission were met in 40% (31-50%) and 29% (23-36%) of patients, respectively. Thirty-two percent of the patients had at least one adverse event, the most commonly reported being mild infection (13%) and worsening of UC, requiring colectomy (13%). A third of the patients (35%) discontinued tofacitinib, most frequently due to primary non-response (51%).

**Conclusions:** Tofacitinib is a safe and effective therapy in real-world UC patients, as previously reported by clinical trials.

**WHAT YOU NEED TO KNOW:** *Background:* Tofacitinib is the first Janus kinase inhibitor (JAKi) approved for the treatment of moderate to severe ulcerative colitis (UC). Real-world data is needed to understand clinical effectiveness of new therapies.

*Findings:* Clinical response rates were consistent with those reported by OCTAVE Induction trials, showing clinical efficacy similar in real-world and controlled clinical trial cohorts. There were no major safety concerns reported.

*Implications for patient care:* The results of this meta-analysis confirm the effectiveness and acceptable safety profile of tofacitinib in a real-world, highly refractory UC population, and further support its use in clinical practice.

## INTRODUCTION

The range of therapeutic agents for inflammatory bowel disease (IBD) management is rapidly expanding. Tofacitinib, a pan-Janus-kinase inhibitor (JAKi) is the first of the new, small molecule drugs to be released for the management of moderate to severe UC. Compared to monoclonal antibodies directed against specific cytokines, JAKi target multiple pathways suggested to modulate intestinal inflammation in UC, such as IL-6, IL-12, IL-15, IL-23 and IFN-γ^1^.

First evidence for tofacitinib efficacy as induction therapy in moderate to severe UC was reported in an 8-week double-blind, placebo-controlled phase 2 trial^2^. This was a dose-defining study in 194 patients that had as primary outcome clinical response at 8 weeks (defined as a decrease in total Mayo score by at least 3 points from baseline and a relative decrease by at least 30%, a decrease in the rectal bleeding subscore of at least 1 point or an absolute rectal bleeding score of 0 or 1). This was achieved by 78% of patients on a 15 mg bd dose, vs. 42% receiving placebo. The majority of secondary outcomes including clinical remission at 8 weeks (Mayo score ≤2, no subscore >1) and endoscopic remission (Mayo endoscopic subscore of 0) were achieved in a greater proportion by patients on a 10 mg dose.

This was followed by two induction (OCTAVE- 1 and −2) and one maintenance (OCTAVE Sustain) phase 3 registration studies, comprising a total of 1139 patients. OCTAVE 1 and 2 demonstrated both efficacy and safety in achieving clinical remission at 8 weeks (defined as a total Mayo score ≤2, no subscore >1 and a rectal bleeding score of 0) by 18.2% and 16.6% of patients on a 10 mg dose, vs. 8.2% and 3.6% in the placebo group. In OCTAVE Sustain, the primary endpoint (clinical remission at 52 weeks) was met in 34.3% and 40.6% of patients on a 5 mg and 10 mg dose, respectively, as compared to 11.1% in the placebo group. In addition, data from an open-label extension of the OCTAVE study (OCTAVE Open) offered evidence-based support for dose escalation after disease flare and showed the possibility of recapturing response after therapy discontinuation^3^, as well as providing longer term safety data.

Rigorously controlled, randomised clinical trials (RCTs) are mandatory to assess treatment efficacy. Importantly however, a number of patients seen in routine clinical practice would not meet inclusion criteria for these studies, particularly those who are older, with refractory disease or significant comorbidity^4^. Indeed, a retrospective analysis of landmark IBD RCT enrollment suggests that only 34% of CD patients and 26% of UC “real-world” patients would have been eligible for inclusion^5^. Common reasons for RCT ineligibility in UC trials were use of topical rectal therapy, being immunomodulator naive, patients with new diagnoses or needing colectomy due to age, comorbidity or concomitant advanced dysplasia^6^.

Uncontrolled, observational data derived outside clinical trial research are often described as real-world evidence (RWE). They collect and use information from daily clinical practice, such as electronic health records, patient registries and surveys or digital health data. By studying a population more representative of clinical practice, RWE may offer additional insights into predictors of effectiveness, drug positioning and reveal unappreciated safety signals, particularly infection and malignancy^7^.

In this systematic review and meta-analysis we aimed to summarise available data on the effectiveness and safety of tofacitinib, derived from early RWE in moderate to severe UC.

## METHODS

### Data sources and searches

This study was conducted according to the PRISMA statement for reporting systematic reviews and meta-analyses (Figure 1)^8^. A systematic search of MEDLINE (via PubMed), Embase, Scopus, Web of Science and Cochrane library was performed. We aimed to identify records that reported real-world experience with tofacitinib from the 30^th^ of May 2018, date of drug approval until the 24^th^ of January 2021. The search strategy included the term “ulcerative colitis” in combination with “tofacitinib” OR “JAK inhibitors”, AND/OR “real-world”. The fields “title, abstract, keyword” and “all fields” were used alternatively. Restrictions for non-English language studies and paediatric population were applied. Additional systematic search through the references of included records was performed to identify articles potentially missed by the search.

**Figure 1.**
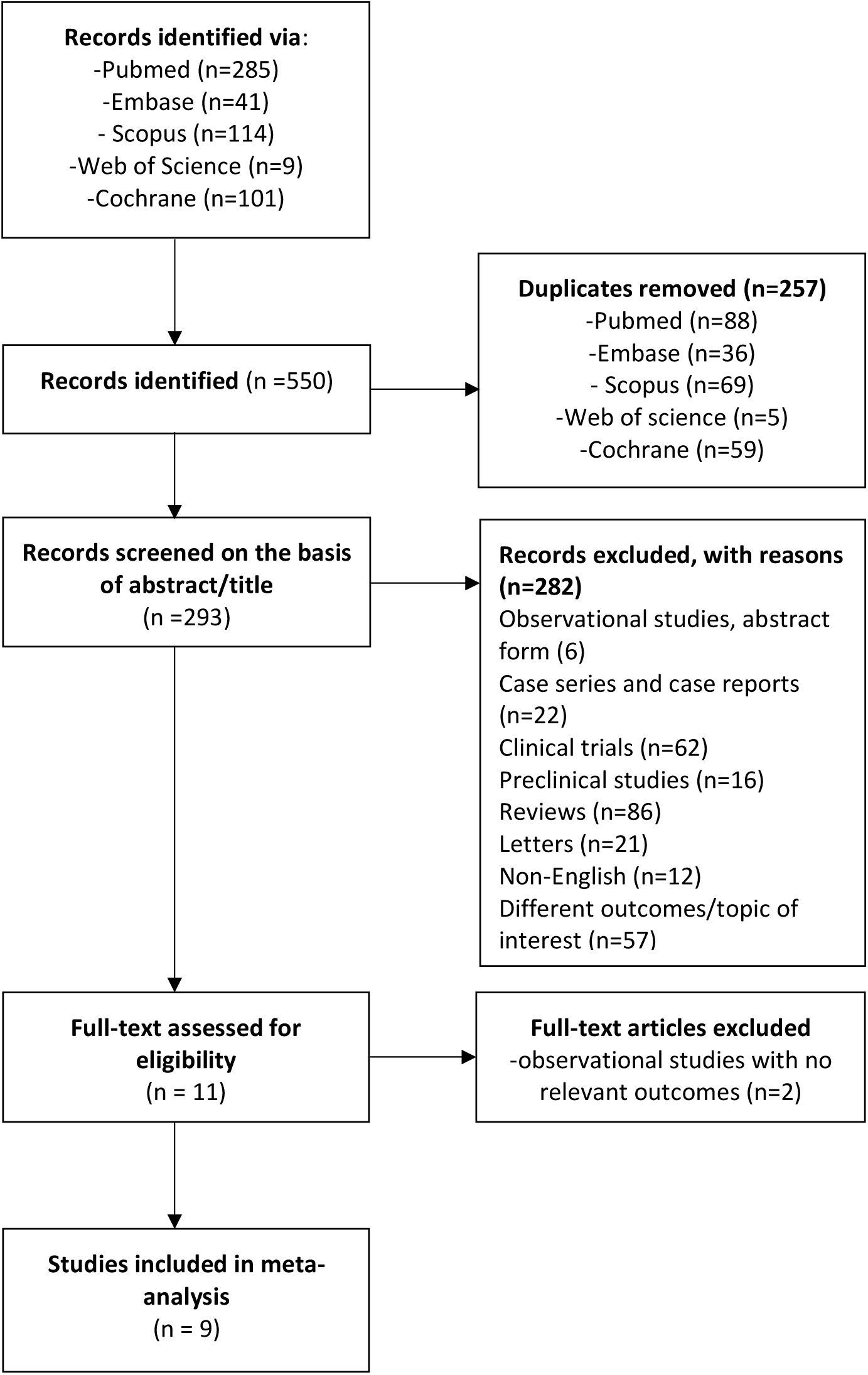
Flow diagram showing results of literature search and study selection

### Study selection and eligibility criteria

After the systematic import of all references into a reference management software (Mendeley), all duplicates were removed. The remaining records were screened for eligibility based on the title and abstract. Only studies that were published in complete form in peer-reviewed literature were considered for the pooled analysis of effectiveness and safety.

We included records that fulfilled the following criteria: (1) study-type - observational studies that reported the effectiveness of tofacitinib in clinical practice; (2) population - adult patients with a diagnosis of UC, with active disease; (3) outcomes - the proportion of patients treated with tofacitinib achieving at least one of the following: clinical response, clinical remission, steroid free remission, endoscopic remission, (4) minimum follow-up - 24 weeks.

For topics that were covered in narrative form, data derived from conference abstracts presented at the European Crohn’s and Colitis Organisation (ECCO), United European Gastroenterology Week (UEGW), Digestive Disease Week (DDW), British Society of Gastroenterology (BSG) and American College of Gastroenterology (ACG) were referenced. Furthermore, we have excluded records classified as case series and case reports.

### Data extraction

The outcomes of interest were extracted by two independent authors (LL and NCC), and disagreements were resolved by consensus. Effectiveness data were grouped under two different temporal phases, according to the time point reported by each study: ‘induction’, which covered outcomes reported at weeks 8, 12 and 14, and ‘maintenance’, which included the outcomes reported at weeks 16, 24 and 26. Data on remission at week 52 was reported only in two studies, therefore it was not included in the analysis. Where both outcomes for weeks 16 and 26 were available, we have included the furthest follow-up time-point for maintenance data. Safety outcomes included the overall number of adverse events, namely infections, venous-thromboembolism, malignancies, need for colectomy and alteration of the lipid profile.

Study-related descriptors included the name of the first author, study design, the number of participants and the median follow-up. Patient demographics that included median age, gender, disease duration (years) and the proportion of patients with extensive disease were collected, as well as disease-related variables (disease activity scores, median levels of faecal calprotectin (FCAL) and CRP), and treatment-related variables, such as tofacitinib dose at induction, bio-naïve status, previous biologics, concomitant steroids and immunosuppressants.

### Descriptors of outcomes

Outcome effectiveness definitions are summarised in Table 1. Established clinical scores such as Partial Mayo Score (PMS) and Simple Clinical Colitis Activity Index (SCCAI) were used to assess clinical response and remission, and Mayo endoscopic subscore to assess endoscopic response and remission.

**Table 1.** Descriptors of tofacitinib efficacy outcomes across real-world studies.

|  |  |
| --- | --- |
| Clinical response | Decrease of $\geq 2$ points<sup>38,39</sup>, $\geq 3$ PMS/ decrease $>30\%$ from baseline<sup>40,20,24</sup><br>Decrease $\geq 1$ point on RBS<sup>38</sup>/absolute RBS=0/1 from baseline<sup>36,20</sup><br>Reduction in SCCAI of $\geq 3$<sup>23</sup><br>Symptomatic improvement but not resolution<sup>22</sup><br>$>50\%$ reduction of symptoms |
| Clinical remission | PMS $\leq 1$, $\leq 2$<sup>20</sup>, without individual subscores $>1$<sup>38</sup><br>PMS$<3$ with combined stool frequency and rectal bleeding subscore $<1$<sup>36</sup><br>SCCAI $<3$<br>Complete resolution of clinical symptoms<sup>22</sup> |
| Steroid-free remission | PMS $\leq 2$ without concomitant use of any steroid preparation<sup>39</sup> |
| Endoscopic remission/<br>Mucosal healing | Mayo endoscopic subscore = 0<sup>24</sup>, 0-1<sup>39</sup> or absence of erosions/ulcerations within 6 months of treatment initiation<sup>36</sup> |
| Relapse | Recurrence of symptoms after an initial response that require therapeutic change<sup>36, 22</sup><br>Worsening of symptoms with endoscopic, radiological or serological (CRP or FCAL) evidence of inflammation that led to escalation/change in medication<sup>20</sup> |
| Failure | Interruption of tofacitinib before the end of follow-up<sup>36</sup><br>Tofacitinib discontinuation due to symptom recurrence in patients in remission, non-response to tofacitinib treatment and serious adverse events<sup>38</sup> |

PMS, partial Mayo score. RBS, rectal bleeding subscale. SCCAI, Simple Clinical Colitis Activity Index. PNR, Primary non-response. LOR, Loss of response. UCEIS, Ulcerative Colitis Endoscopic Index

### Quality assessment of included studies

Two authors (NP and LL) independently assessed the quality of included studies using the Newcastle Ottawa scale for cohort studies^9^ (Supplementary file. Newcastle-Ottawa scale) (Supplementary table 1). The following criteria were evaluated: selection (UC patients with moderate to severe disease and lack of / poor response / intolerance to biologics that received treatment with tofacitinib for at least 8 weeks) and assessment of outcomes. All studies are uncontrolled cohort studies, therefore the domain “comparability” and “selection item 2”, describing the non-exposed cohort, were not applicable for this meta-analysis. As a result, the maximum score achievable by any of the studies reported was 7 instead of 9. Newcastle-Ottawa scores were originally defined as high (score 7-9), moderate (score 4-6) or low (score 0-3). We have graded as “high quality” all studies that fulfilled a score of 6 or 7, and as “moderate quality” the studies with a score of 4 or 5.

### Statistical analysis

The R packages meta and metafor^10, 11, 12^ were used to calculate pooled proportions of patients, alongside 95% confidence intervals, responding to tofacitinib at induction and maintenance specified timelines. Pooled proportions of patients who experienced an adverse event or endoscopic remission across the entire study durations were also calculated. Due to the relatively small number of participants in the studies, logit transformations were applied before fitting random effects models using the DerSimonian and Laird method^13^. Random effects models were used as the cohort populations were not consistent between studies.

At least four studies were required for each analysis. Outliers were detected using externally standardised residuals and leave-one-out residual heterogeneity. Influential studies were found by investigating the influence deleting a study had on each individual parameter estimate using four different measures. If at least one measure found a study to be influential, the study was overall deemed to be influential. Detailed explanations of the methods used to detect outlier and influential studies can be found in the literature^14^. Evidence of publication bias was sought using funnel plots, and formally tested for using Egger’s regression test with a significance level of 5%^15^. R version 4.0.3 was used for this analysis. R code is available to reproduce the analysis at https://github.com/nathansam/tofameta. Heterogeneity was assessed using I2 values (the percentage of variation across the studies due to heterogeneity)^16^.

## RESULTS

### Outcomes of search strategy

The search strategy identified 550 studies via Pubmed, Embase, Scopus, Web of Science and Cochrane databases (Figure 1). After removing the duplicates, 293 studies were assessed against the predefined inclusion criteria. Of these, studies that did not meet the criteria for definition of outcomes or addressed different research topics, other publication types (RCTs, preclinical studies, reviews/meta-analyses, case series and case reports, comments, editorials, published erratum and letters) (n=282) were excluded (Figure 1). Two observational studies that did not fulfil the criteria on minimum follow-up duration or reported outcomes were furthermore excluded. Nine studies, 7 retrospective and 2 prospective, were therefore combined for qualitative analysis. The mean Newcastle-Ottawa score among the 9 included studies was 6, with 8 / 9 studies demonstrating high quality scores (Supplementary Table 1).

### Characteristics of patients across studies

A total of 830 UC patients were included in the meta-analysis, with a median age of 40 years (IQR 37-45) and a median disease duration of 7 years (IQR 5.2-9.4) (Supplementary table 2). Patients were followed up for a median duration of 31 weeks (23.5-41.5). The proportion of patients with extensive disease (Montreal classification E3) was 55% (454/830). At baseline, the median PMS was 6 (IQR 5-6), CRP 4.5mg/L (IQR 2-7) and FC 1265 mcg/g (IQR 674-1898). Eighty-one percent of patients (674/830) were previously treated with anti-TNF antagonists alone, whereas 57% (452/800) of patients across 8 studies had failed vedolizumab as second-line therapy. An additional 5% (28/534) had previously been treated with ustekinumab, as reported by 4 studies. Half of the patients (412/830) were undergoing concomitant therapy with systemic corticosteroids, whereas concurrent immunomodulators use weas reported in 8% of patients. Tofacitinib use in biologic naive population was quantified in 2 studies, comprising 69 of 257 (59%) patients^17^. Tofacitinib dose was either 5 or 10 mg twice daily, according to investigator’s protocol (Supplementary table 2).

### Efficacy outcomes

Outcomes were heterogeneously reported across studies, with many studies reporting the outcomes at different timepoints. We have pooled the results from the number of studies that had analysed the outcome of interest, therefore the number of patients per outcome assessment may differ.

### Induction of clinical response and remission

Clinical response and remission were reported in 496 patients across 6 studies (Table 2). Median time to outcome assessment in the induction group was 8 weeks (IQR 8-12). Clinical response was achieved in 51% (262/496, 95% CI 0.41-0.60%) and clinical remission in 37% (187/496, 95% CI 30-45%) of patients respectively (Figure 2a, b). Steroid-free remission was 32% (129/391, 95% CI 25-40%), as assessed by 5 records (Supplementary figure 1a).

**Table 2.**
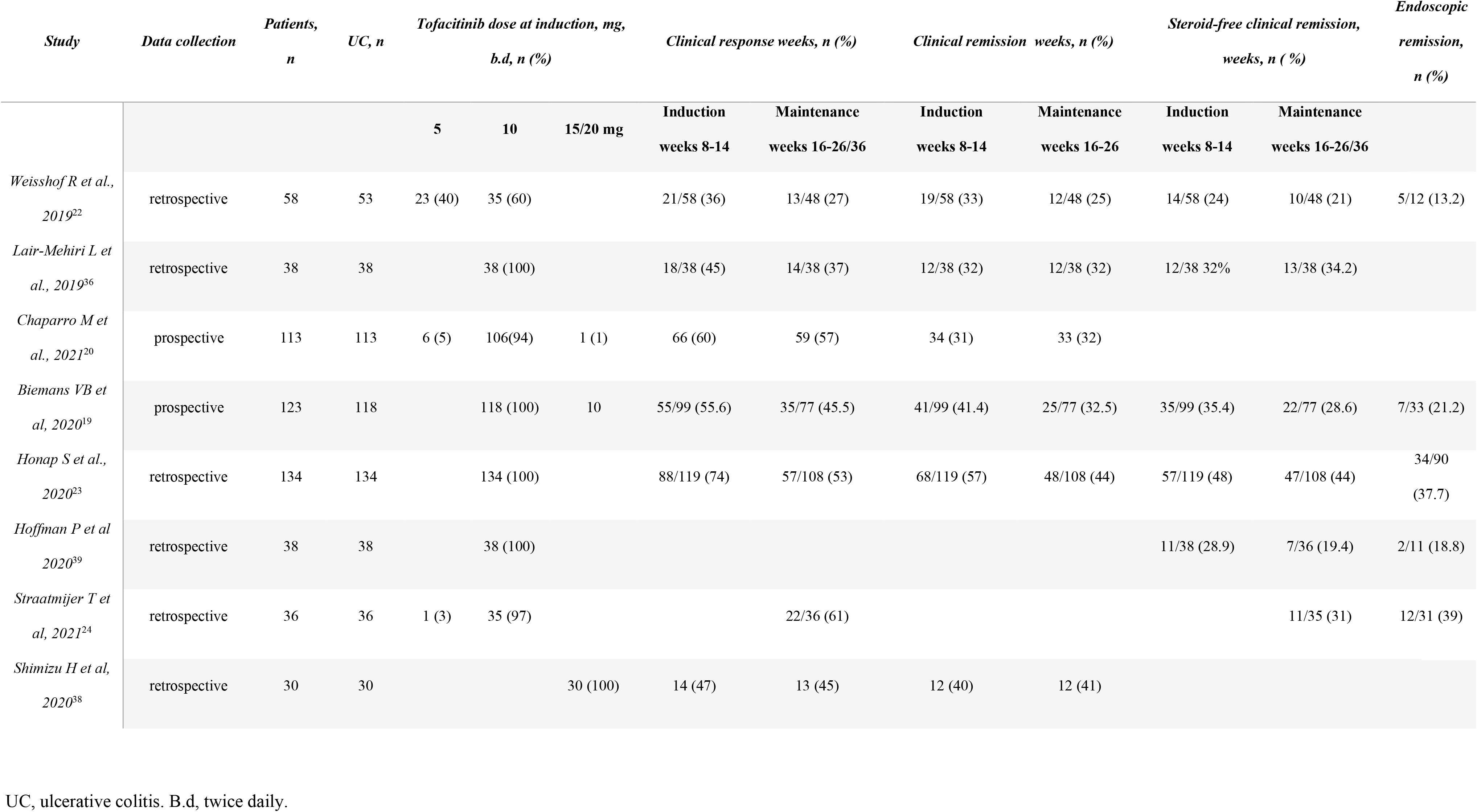
Efficacy of tofacitinib treatment across real-world studies in ulcerative colitis patients at induction (8,12,14) and maintenance (16, 24, 26) timepoints.

**Figure 2.**
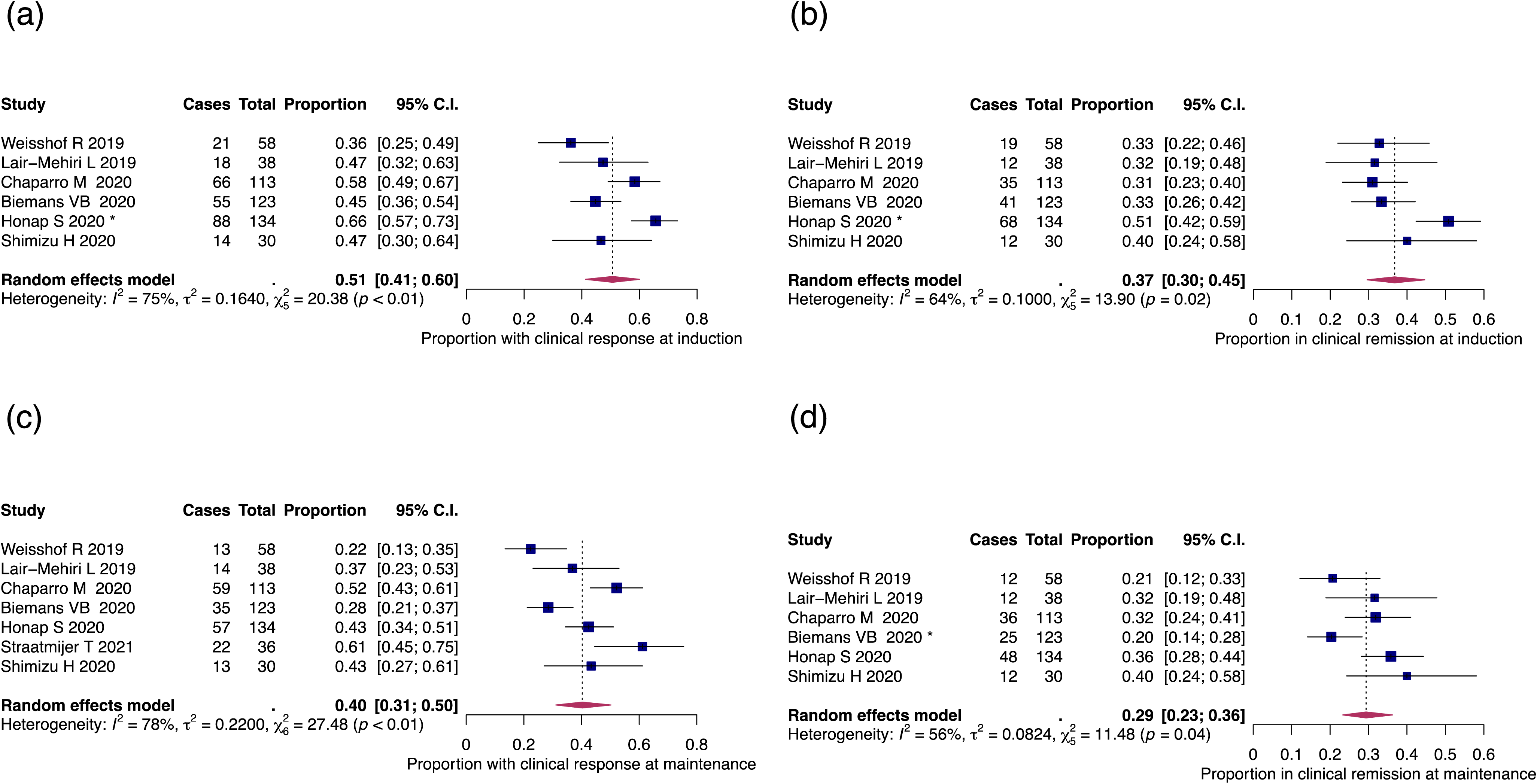
Pooled efficacy of tofacitinib in ulcerative colitis (UC) real-world patients: a) clinical response during induction (8, 12, 14 weeks); b) clinical remission during induction (8,12,14 weeks); c) clinical response during maintenance (16, 24, 26 weeks); d) clinical remission during maintenance (16, 24, 26 weeks). Influential studies are denoted with *

**Figure 3.**
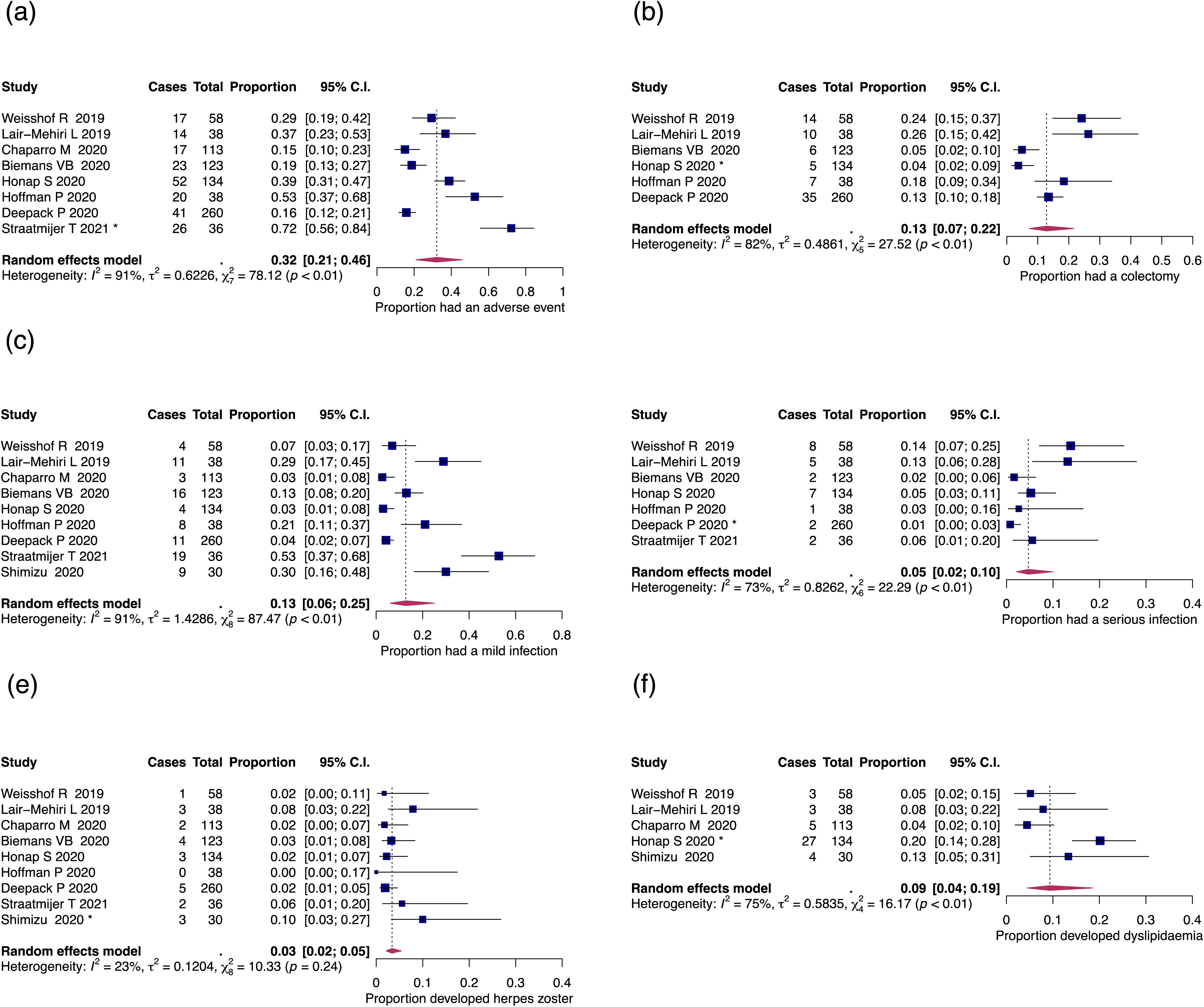
Adverse events among real-world UC patients. a) Proportion of patients that had at least one adverse event; b) colectomy; c) mild or moderate infection; d) serious infection; e) herpes zoster infection; f) dyslipidaemia. Influential studies are denoted with *

### Maintenance of clinical response and remission

Median time to outcome assessment in the maintenance group was 24 weeks (IQR 18-24) (Table 2). Clinical response and remission in the maintenance phase were reported in 532 and 496 patients across 7 and 6 studies, respectively. Clinical response was achieved in 40% (213/532, 95% CI 31-50%) and clinical remission in 29% (145/496, 95% CI 23-36%) of patients respectively (Figure 2c, d). The proportion of patients in steroid-free remission was 25% (109/427, 95% CI 18-33%), as reported by 6 studies (Supplementary figure 1b). Endoscopic evaluation was conducted at the end of follow-up in 60 patients across 5 studies, and endoscopic remission was achieved by 34% (60/177, 95% CI 26-43%) of patients (Supplementary figure 2).

### Predictors of response and remission

Due to significant heterogeneity in reporting patients’ characteristics and outcomes, it was not possible to perform pooled analysis for predictors of response. There were fewer than 10 studies with case data for any outcome of interest. As a result, we lacked the statistical power required to perform valid subgroup analyses^18^.

Factors independently associated with clinical response at week 8 were reported in 1 study^17^. Multivariable analysis in this work revealed that patient’s treatment naive status (OR 4.50, 95% CI 1.64-12.37) and a higher albumin (OR 2.63, 95% CI 1.02-6.80) were associated with a greater chance of achieving clinical response at week 8, whereas the concomitant use of corticosteroids at the start of tofacitinib treatment (OR 0.22, 95% CI 0.08-0.58), male gender (adjusted hazard ratio 0.25, 95% CI 0.08-0.83) and pancolitis were associated with a lower chance of achieving week 8 response^17^. In the same study, prior exposure to two biologic classes had lower rates (16.7%) of endoscopic healing at 6 months compared to bio-naive patients (87.1%) and those with exposure to one biologic agent (57.1%). A similar finding was reported by a Dutch prospective observational registry, which showed that steroid-free clinical remission rate at week 24 was influenced by prior exposure to vedolizumab in a multivariable analysis (OR 0.301, 95% CI 0.100–0.907)^19^.

Clinical predictors, such as a higher PMS at week 4 was the only variable associated with the likelihood of achieving short term remission at week 8 (OR 0.2; 95% CI 0.1 –0.4), but not at week 16, as reported by Chaparro et al.^20^. The multivariable analysis by Verstockt et al.^21^ showed that a higher baseline albumin and a lower Mayo endoscopic sub-score were independent predictors of endoscopic (OR 1.06 and 0.59, respectively) and biological remission (OR 1.06 and 0.57, respectively). Tofacitinib dose did not correlate with clinical response at week 8^22^.

### Treatment discontinuation

Treatment discontinuation was reported in 35% of patients (254/800, 95% CI 28-43%) across 8 studies (Supplementary figure 3a). The most common reason for discontinuation was PNR, reported in 51% of these patients (111/194, 95% CI 34-68%) (Supplementary figure 3b). Other reasons for treatment discontinuation were adverse events (AE) in 20% of patients (44/241, 95% CI 13-30%) (Supplementary figure 3c), need for colectomy 19 % (29/147, 95% CI 6-47%) and patient request (0.8%, 3/370) (Table 3). Median time to discontinuation of therapy was 9 weeks19, 23, 24.

**Table 3.** Adverse events of tofacitinib treatment and therapy discontinuation in ulcerative colitis patients across real world studies.

| Study | Patients,<br>n | Patients<br>with AE, n | Herpes<br>zoster, n | Infection, n |  |  | VTE, n | Dislipidemia,<br>n | Malignancy,<br>n | Colectomy n,<br>% | Therapy<br>discontinuation, n (%) | Reasons for therapy discontinuation, n (%) |  |  |  |  |
| --- | --- | --- | --- | --- | --- | --- | --- | --- | --- | --- | --- | --- | --- | --- | --- | --- |
|  |  |  |  | Total | mild/<br>moderate | serious |  |  |  |  |  | AE | Colectomy | PNR | LOR | Patient's<br>choice |
| Weisshof R et al., 2019<sup>22</sup> | 58 | 17 | 1 | 12 | 4 | 8 | 0 | 3 |  | 14 (24.1) | 26 (43.1) | 5/26 (19.2) |  | 6/26 (23.7) |  |  |
| Lair-Mehiri L et al., 2019<sup>36</sup> | 38 | 14 | 3 | 16 | 11 | 5 | 0 | 3 |  | 10 (26) | 16 (42) | 5/16 (31.2) | 7/16 (43.7) | 4/16 (25) |  |  |
| Chaparro M et al., 2021<sup>20</sup> | 113 | 17 | 2 | 3 | 3 |  | 0 | 5 | 1 |  | 45 (40) | 7 /45 (15.5) |  | 29/45 (64.4) | 8/45 (17.7) | 1/45 (2.2) |
| Biemans VB et al, 2020<sup>19</sup> | 123 | 23 | 4 | 16 | 16 | 0 | 0 |  |  | 6 (4.9) | 46 (37.4 ) | 7/46 (15.2) |  | 35/46 (76.1) | 3/46 (6.5) | 1/46 (2.2) |
| Honap S et al., 2020<sup>23</sup> | 134 | 52 | 3 | 11 | 4 | 7 | 0 | 27 |  | 5/134 (3.7) | 48 (35.8) | 21/48(43.7) |  | 31/48 (64.5) | 14/48 (29.1) | 1/48 (2.0) |
| Hoffman P et al, 2020<sup>39</sup> | 38 | 20 | 0 | 9 | 8 | 1 | 0 |  |  | 7 (18.4) | 13(34.2) |  | 1/13 (7.6) |  | 13/13 (30.7) |  |
| Deepack P et al, 2020<sup>25</sup> | 260 | 41 | 5 | 13 | 11 | 2 | 2 |  | 2 | 35(13) | 47 (18) | 12/47 (25.5) | 1/47 (2.1) |  | 34/47 (72.3) |  |
| Straatmijer T et al, 2021<sup>24</sup> | 36 | 26 | 2 | 21 | 19 | 2 |  |  |  |  | 14/36 (33) | 6/36(16.6) |  | 6/36 (16.6) |  |  |
| Shimizu H et al, 2020<sup>38</sup> | 30 |  | 3 | 9 | 9 | 0 |  | 4 |  |  |  |  |  |  | 16 (53.3) |  |

AE, Adverse events. IQR, interquartile range. VTE, venous thromboembolism. PNR, primary non-response. LOR, loss of response

### Predictors of treatment discontinuation and PNR

The only factor associated with treatment discontinuation was higher PMS at week 8 in a multivariable analysis^20^. A prospective cohort following endoscopic and histologic outcomes of tofacitinib treatment in 35 patients refractory to anti-TNF and vedolizumab^21^ reported that 10 of the patients who experienced PNR to one anti-TNF discontinued tofacitinib due to same reason. Moreover, PNR to two anti-TNFs resulted in PNR to tofacitinib in 4 out of 5 patients. In contrast, for the 8 patients previously treated with vedolizumab, only 3 were nonresponders to tofacitinib, as reported by same study. In the work of Honap et at al^23^, younger age at treatment initiation (median of 28 years, IQR 23.0-37.5, aHR 1.04 (95% CI 1.01-1.07)) and elevated CRP at baseline (aHR 0.29 (95% CI 0.12-0.66), for every ten-fold increase in baseline CRP) were independently associated with PNR at week 8. In addition, response and remission rates were not different stratified by prior biologic use, as reported by the same study. These findings were confirmed by other reports^22^, that have not detected a link between PNR and previous biologic therapy.

### Safety outcomes

Relevant safety outcomes across all studies are summarised in Table 3. The proportion of patients that experienced at least one AE was 32% (210/800, 95% CI 21-46%), during a median follow-up of 18 weeks (IQR 11-42), as reported across 8 studies (Figure 4a). The most common AEs were mild infection 13% (47/830, 95% CI 6-25%), and colectomy 13% (77/651 95% CI 7-22%); other AEs were dyslipidaemia 9% (42/373, 95% CI 4-19%), serious infection leading to hospitalisation 5% (27/687, 95% CI 2-10%), and herpes zoster infection 3% (23/830 95% CI 2-5%) (Figure 4 b-f). Only two episode of deep vein thrombosis^25^ and one case of malignancy (excluding non-melanoma skin cancer)^20^ were reported.

## DISCUSSION

Tofacitinib is the first small molecule JAKi licensed for moderate to severe UC. In this meta-analysis we have summarised the RWE for tofacitinib use in UC.

Our meta-analysis showed clinical response induction rates (51%) comparable with those reported from OCTAVE Induction trials 1 and 2 (59.9% and 55%, respectively). Clinical remission at 8 weeks were divergent: 37% across real-world cohorts vs 18.5% and 16.8% for OCTAVE Induction 1 and 2, respectively. Whilst maintenance of clinical response at either 16, 24 and 26 weeks from our RWE analysis was lower than those in OCTAVE Sustain (40.0% versus 63.6%), clinical remission rates at the same time points were broadly comparable (29.0% versus 34.3%) at 24 weeks in OCTAVE Sustain. Testing for differences in outcomes between OCTAVE and the observational studies was also not possible due to differences in outcome definitions.

There are several factors to be considered when comparing our data with results from clinical trials. Firstly, the timing of efficacy evaluation varied widely between studies. Therefore, we pooled the results into induction (8-14 weeks) and maintenance (16-26 weeks) timepoints since most studies reported data points at 8 and 24 weeks. Secondly, there was heterogeneity among studies in reporting clinical information regarding patients’ characteristics; therefore multivariable analysis to identify predictors of response and remission could not be performed.

The majority of RWE patients (81%) were refractory to 1 or 2 anti-TNF agents, whereas 57% and 5% had previously been exposed to vedolizumab and ustekinumab. In one of the two studies that reported tofacitinib effectiveness in biologic naive patients, clinical and endoscopic response rates were higher compared to those previously treated with biologics^17^.

Overall, tofacitinib was well tolerated among patients and had a comparable safety profile with data from clinical trials. In our meta-analysis, the proportion of patients that experienced at least one adverse event was lower (32%) in comparison to 56% and 54% reported by OCTAVE induction trials; this is likely an artefact from the formal RCT AE reporting process. Furthermore, we reported lower rates for infection (mild/moderate 13%, serious infection 5% vs 23% and 18%, respectively) but higher herpes zoster infection rates (4% herpes vs 0.6 and 0.5%).

JAK inhibition has also been associated with alteration of serum lipid profile, the occurrence of major cardiovascular adverse events (MACE) and venous thromboembolism (VTE). Our pooled analysis showed a 9% rate of dyslipidaemia among 5 studies, 2 occurrence of deep vein thrombosis^25^ and no major cardiovascular adverse events reported among real-world patients. A post-hoc analysis of OCTAVE clinical trials comprising 1157 UC^26^ patients reported one occurrence of deep vein thrombosis and 4 pulmonary emboli in the overall cohort; all were treated with tofacitinib 10 mg twice daily and had other risk factors for VTE. With regards to MACE, such as haemorrhagic stroke, aortic dissection, acute coronary syndrome and myocardial infarction the IR was 0.2 (0.1-0.6%) based on 4 patients in the overall cohort.

There was one case of malignancy (excluding non-melanoma skin cancer) in our cohort of real-world patients^20^. An integrated safety analysis^27^ based on all RCTs of tofacitinib in UC (phase II induction^2^, OCTAVE Induction 1, 2 and Sustain^28^, OCTAVE Open^29^) reported similarly low rates of malignancies, with an IR of malignancies (excluding non-melanoma skin-cancer) of 0.7% (95% CI 0.3-1.2) for 11/1157 patients.

Treatment discontinuation rate across RWE studies was 35%, mostly due to PNR (51%) and AE (20%). Other reasons for treatment discontinuation were need for colectomy (19%) and patient’s preference (0.8%). In OCTAVE induction trials treatment discontinuation rate irrespective of the reason was 6.5% (32/492) and 7.5% (33/435) for Induction 1 and 2 trials respectively, whereas in OCTAVE Sustain 39.8% (157/394) discontinued treatment in both 5 and 10 mg dose arms.

It is noteworthy that across our RWE dataset, the median age was 41 years. This should be borne in mind when comparing AEs, especially MACE and malignancy, with data from rheumatology with tofacitinib. Preliminary reports of the ORAL Surveillance (A3921133; NCT02092467) study in rheumatoid arthritis have shown an increase in MACE and malignancy with tofacitinib versus infliximab in patients over 50 years of age with at least one additional cardiovascular risk factor^30^.

There is limited evidence on the use of tofacitinib for moderately-to-severe UC outside RCTs. To safeguard our findings against the emergence of future publications, we repeated our analyses with the inclusion of conference abstracts that have not to date been published in full^31,17,21,32,33,34^. This search had yielded an additional 6 studies including a total of 332 patients on top of those reported here. The results from this analysis did not deviate substantially from the findings we have reported (data not shown). On the contrary, the inclusion of abstracts was shown to reduce the number of outcomes with significant publishing bias from four to two. Sensitivity analysis of full papers alone and both full papers and abstracts included showed that overall, the inclusion of abstracts augmented our findings by contributing to increased precision and comprehensiveness of the analysis (Supplementary figure 4).

Estimates of heterogeneity calculated using I^2^ for many of our outcomes was greater than 75% which is commonly interpreted to indicate considerable heterogeneity. However, for many of the outcomes of interest, there was a limited number of events or studies and as such, I^2^ is arguably not reliable in the context of this meta-analysis^35^. This was most notable when investigating treatment discontinuation due to AE which yielded a lower bound of 0 and an upper bound of 94.3% for a 95% confidence interval of I^2^.

This meta-analysis has two major limitations: firstly, the majority of studies are retrospective, leading to significant heterogeneity in study population, data collection, definitions of outcomes, tofacitinib dose, time of drug exposure, length of follow-up, and endoscopic evaluation. Therefore, extracting data for predictors of response or testing for differences in outcomes between real-world patients and the OCTAVE trials was not achievable. Secondly, real-world studies comprising patients treated before the 30^th^ of May 2018^22, 36, 33^ used off-label medication or accessed it through compassionate programs which restricted the selection criteria to severe, refractory cases. Other studies^37^ exclusively included patients refractory to other biologics (anti-TNF and vedolizumab).

To the best of our knowledge, this is the first study to have summarised available efficacy and safety evidence in the real-world population with UC, across exclusively peer-reviewed data. Tofacitinib appears to be at least as effective as it was previously demonstrated in clinical trials, with an attractive safety profile. Long-term follow-up data may provide additional information with regards to non-response, effectiveness in special populations (pregnancy, children, elderly) and adverse events (malignancies).

## Data Availability

R code and extracted data will be made available to reproduce the meta-analysis at https://github.com/nathansam/tofameta upon acceptance by a journal.

## Abbreviations

AE: adverse events
aHR: adjusted hazard ratio
CI: confidence interval
b.d: bi-daily
IBD: inflammatory bowel disease
IL: interleukin
IQR: inter-quartile range
JAKi: Janus-kinase inhibitor
LOR: loss of response
MACE: major adverse cardiovascular events
OR: odds ratio
PMS: partial Mayo score
PNR: primary non-response
UC: ulcerative colitis
UCEIS: ulcerative colitis endoscopic index of severity
RCT: randomized controlled trial
RWE: real-world evidence
RBS: rectal bleeding scale
SCCAI: Simple Clinical Colitis Activity Index
VTE: venous thromboembolism.

---

**Supplementary figure 1.**
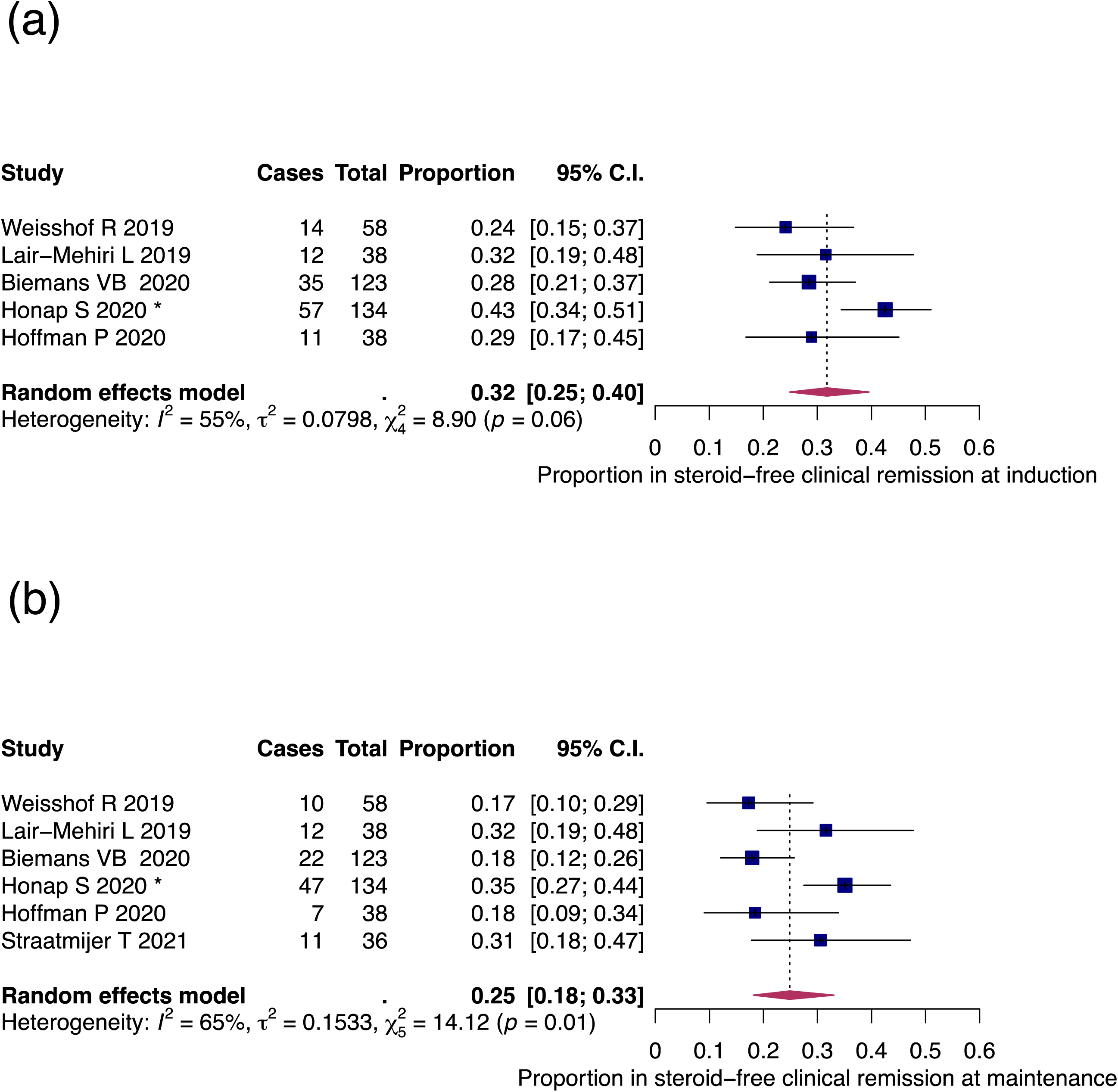
Pooled efficacy of tofacitinib in UC real-world patients: steroid-free clinical remission. a) Steroid-free clinical remission during induction (8, 12, 14 weeks); b) steroid-free clinical remission during maintenance (16, 24, 26 weeks). Influential studies are denoted with *

**Supplementary figure 2.**
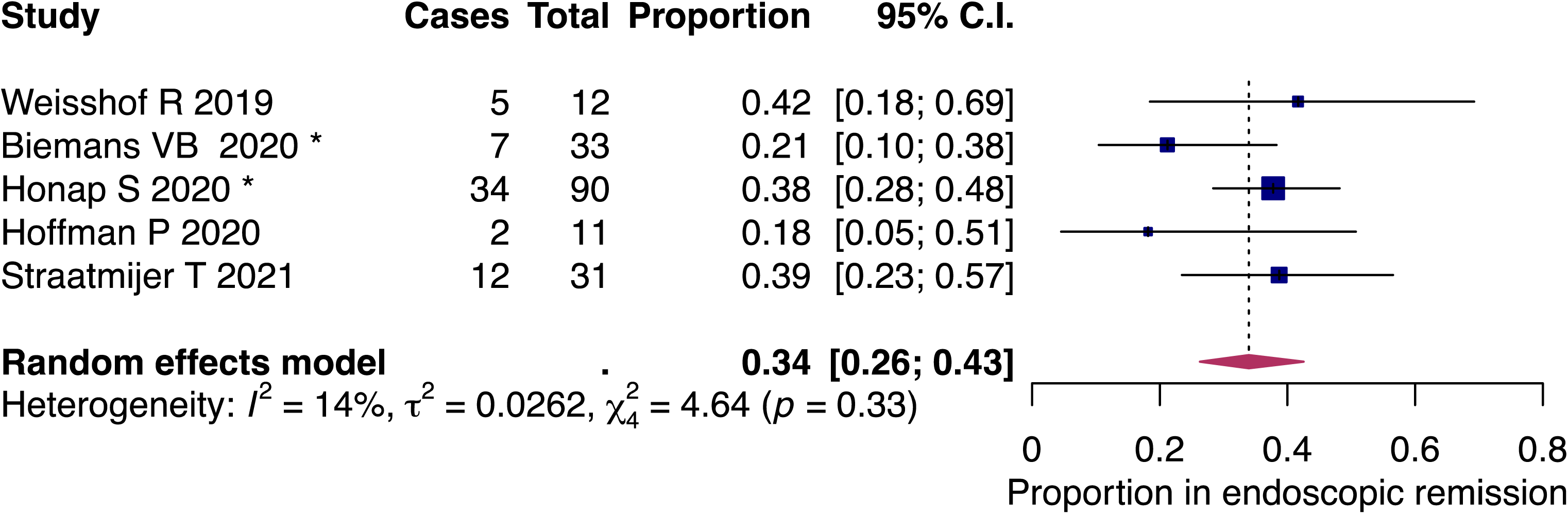
Pooled efficacy of tofacitinib in UC real-world patients: endoscopic remission. Influential studies are denoted with *

**Supplementary figure 3.**
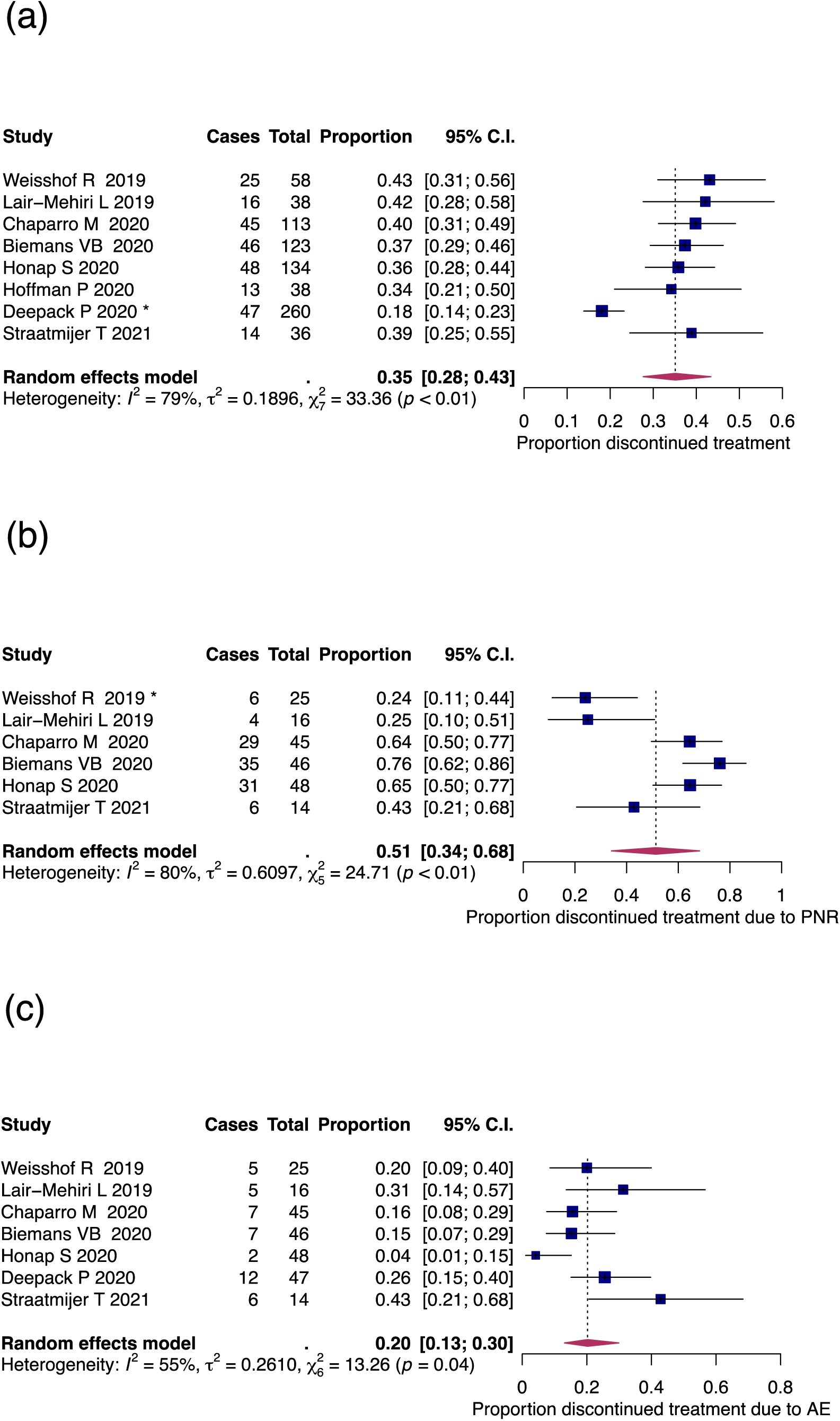
Tofacitinib therapy discontinuation among real-world UC patients according to reason for discontinuation: a) pooled tofacitinib therapy discontinuation; b) discontinuation due to primary non-response (PNR); c) discontinuation due to adverse events (AE). Influential studies are denoted with *

**Supplementary figure 4.**
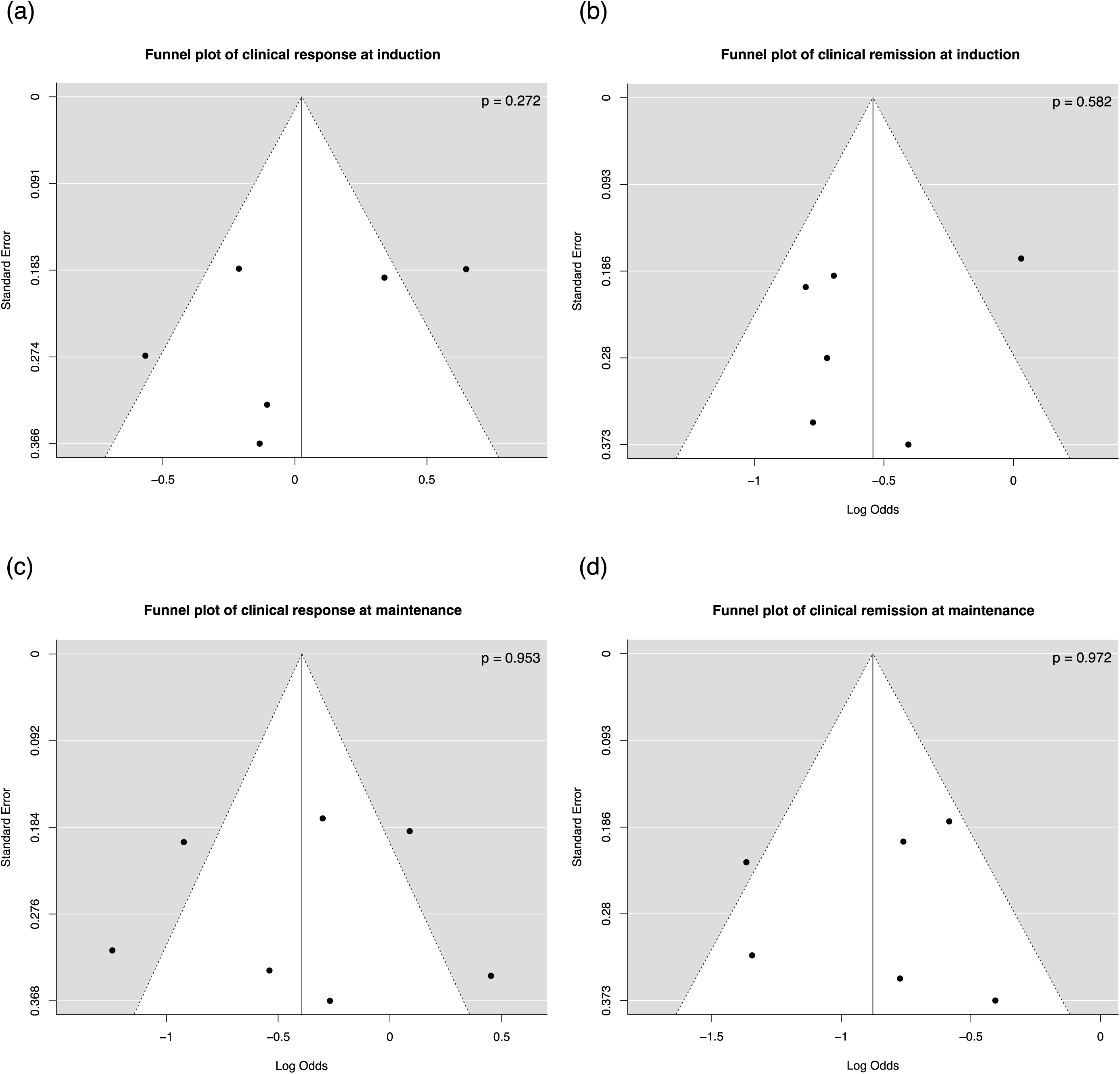
Funnel plots of primary efficacy outcomes with Egger’s regression test p-values. a) Clinical response at induction; b) clinical remission at induction; c) clinical response at maintenance; d) clinical remission at maintenance.

